# Population mobility and the development of Botswana’s generalized HIV epidemic: a network analysis

**DOI:** 10.1101/2023.02.01.23285339

**Authors:** Janet Song, Justin T. Okano, Joan Ponce, Lesego Busang, Khumo Seipone, Eugenio Valdano, Sally Blower

## Abstract

The majority of people with HIV live in sub-Saharan Africa, where HIV epidemics are generalized. For these epidemics to develop, populations need to be mobile. However, population-level mobility has not yet been studied in the context of the development of generalized HIV epidemics. Here we do so by studying historical migration data from Botswana which has one of the most severe generalized HIV epidemics worldwide; in 2021, HIV prevalence was 21%. The country reported its first AIDS case in 1985 when it began to rapidly urbanize. We hypothesize that, during the development of Botswana’s epidemic, the population was highly mobile and there were substantial urban-to-rural and rural-to-urban migratory flows. We test this hypothesis by conducting a network analysis using a historical time series (1981 to 2011) of micro-census data from Botswana. We found 10% of the population moved their residency annually, complex migration networks connected urban with rural areas, and there were very high rates of rural-to-urban migration. Notably, we also found mining towns were both important in-flow and out-flow migration hubs; consequently, there was a very high turnover of residents in towns. Our results support our hypothesis, and together, provide one explanation for the development of Botswana’s generalized epidemic.

## INTRODUCTION

Over 25 million people live with HIV infection in sub-Saharan Africa. All of the HIV epidemics in this continent are generalized: in these type of epidemics, the epidemic is dispersed throughout the country. Therefore, a population needs to be highly mobile in order for a generalized epidemic to have developed. However, although the epidemiology of HIV in sub-Saharan Africa has been widely studied (Farley et al., 2022; Read et al., 2022), the role of population-level mobility patterns in the development of generalized HIV epidemics has not been assessed for any country on the continent. Botswana has one of the most severe HIV epidemics worldwide, and reported its first AIDS case in the early 1980s. Since then, the epidemic has become generalized and hyper-endemic: in 2021, HIV prevalence in adults (15 to 64) was 21% (Mine et al., 2022). Simultaneous to the development of its generalized HIV epidemic, Botswana has undergone rapid urbanization. It was predominantly rural in 1981 (Tarver, 1984; Tarver & Miller, 1987), but had become predominantly urban (Statistics Botswana, 2014) by 2011; by that time Botswana’s HIV epidemic had stabilized (CSO Botswana & NACA, 2009). We hypothesize that – during the development of Botswana’s epidemic – there was a high level of population mobility, and substantial urban-to-rural migratory flows. We test this hypothesis by conducting a network analysis using a historical time series of micro-census (i.e., individual-level) data collected in Botswana. These data contain information both on migration and urbanization. The time series covers the time period from when the epidemic was first apparent to when it stabilized in 2011. We use these data: (i) to estimate the annual incidence (at the national level) of internal migration over three decades (1981 to 2011), (ii) to characterize migrants on the basis of gender and age, (iii) to reconstruct internal migration networks (in order to identify large-scale population movements and connectivity patterns), (iv) to identify migration hubs, and (v) to quantify urban-to-rural and rural-to-urban migratory flows. The Government of Botswana defines internal migration as residents changing their place of permanent residence within their home country. We discuss our results in the context of understanding the development of the generalized hyper-endemic HIV epidemic in Botswana.

Between 1981 and 2011, the population of Botswana increased from just under one million to over two million (Statistics Botswana, 2014) and the country became progressively urbanized. In 1981, the vast majority of the population lived in small rural villages. Only ∼18% of the population were living in urban areas (Statistics Botswana, 2014): either in one of the two cities (Gaborone, the capital, or Francistown) or one of the four towns (Lobatse, Selebi Phikwe, Orapa, and Jwaneng). Lobatse is an administrative center, and the other three are mining towns. Selebi Phikwe is based on copper and nickel mining, and was founded in the early 1970s. Orapa and Jwaneng are based on mining diamonds, and were founded in the late 1960s and early 1980s, respectively. By 2011, only ∼36% of the population were living in rural areas (Statistics Botswana, 2014). The remaining population were living in one of the three types of urban centers: a city (there were still only two cities), a town (there were now five towns following the 1991 addition of Sowa, a mining town for soda ash) or an urban village. In Botswana, an urban village is defined as a settlement with at least 5,000 individuals and 75% of the workforce engaged in non-agricultural economic activities (Statistics Botswana, 2014). The urban villages developed by in situ urbanization, which is defined as rural settlements transforming into urban areas by expanding their non-agricultural activities and increasing economic linkages with neighboring areas (Moriconi-Ebrard et al., 2020). Botswana consists of 28 administrative districts (Figure 2— figure supplement 1) (Okano et al., 2021). Each city and town are separate administrative districts, and the remaining 21 districts contain at least one urban village and many rural villages.

A great deal is known about the epidemiology of HIV in Botswana in terms of the increase in prevalence and geographic variation in prevalence (Magosi et al., 2022; Novitsky et al., 2015; Novitsky et al., 2020; Okano et al., 2021). The first case of AIDS was reported in 1985 from the nickel mining town of Selebi Phikwe (African Natural Resources Center, 2016); in the late 1980s, additional cases were reported from two diamond mining towns (Jwaneng and Orapa) and the city, Francistown. In 1990, the first HIV Sentinel Surveillance Survey of antenatal clinic (ANC) attendees (15 to 49 years old) was conducted in Gaborone and in one rural district (Boteti): HIV prevalence was found to be 6% and 4%, respectively (UNAIDS & World Health Organization, 2004). In 1992, a national sentinel surveillance survey began and was conducted annually to 2011. National prevalence in pregnant women was found to have already reached a very high level (18%) by 1992, and continued to increase for the next eight years: 23% by 1993, 32% by 1995, and 39% by 2000 (UNAIDS & World Health Organization, 2004). Based on the ANC data (UNAIDS & World Health Organization, 2004), HIV prevalence in the cities and the mines was the highest. In Francistown, prevalence was 8% in 1991, 24% by 1992, and 44% by 2000. In Gaborone, prevalence rose from 15% in 1992 to 36% by 2000. In the mining town of Selebi Phikwe, prevalence rose from 27% in 1994 to 50% by 2000. By the late 1990s, a high AIDS morbidity and mortality rate in miners led the diamond mining company Debswana to conduct an HIV prevalence survey of its (male) employees. The survey took place in 1999: 29% of miners were found to be infected with HIV (Barnett et al., 2002). The first population-level survey in Botswana to test participants for HIV (BAIS II, the Botswana AIDS Impact Survey) was conducted in 2004 (NACA & CSO Botswana, 2005). It found that, at that time, prevalence in the general population was ∼29% in women and ∼20% in men aged 15 to 49 years old. Notably, phylogenetic studies have shown that viral lineages have dispersed widely throughout Botswana suggesting large-scale population-level movements have occurred (Magosi et al., 2022; Novitsky et al., 2015; Novitsky et al., 2020).

## RESULTS

### Estimated incidence of internal migration

We estimated the Crude Migration Intensity (CMI) (Bell et al., 2002) for Botswana in the 12 months before each census was conducted (i.e., between 1980 and 1981, 1990 and 1991, 2000 and 2001, and 2010 and 2011). The CMI represents the overall incidence, or level of internal migration, per hundred residents over a specified time interval (Bell et al., 2002); it is an indicator of the propensity of the population to move, and is a measure of migration both between districts and within districts. The CMI remained markedly high and constant: 9.05 per hundred persons between 1980 and 1981, 10.45 per hundred persons between 1990 and 1991, 10.73 per hundred persons between 2000 and 2001, and 10.33 per hundred persons between 2010 and 2011. We found that migrants were more likely to move between districts than to move within districts; this propensity, calculated as a ratio (the number of migrants moving between districts relative to the number of migrants moving within districts), increased over time from 1.25 (1981) to 2.40 (2001 and 2011).

### Age profiles of migrants

Gender-stratified age profiles of migrants are presented in Figure 1. These results show that the type of individual who migrated within Botswana at the time of each census was markedly similar, with respect to gender and age. Approximately 50% of migrants were women, and the most common age to migrate (for both women and men) was between 16 and 20 years old.

**Figure 1:**
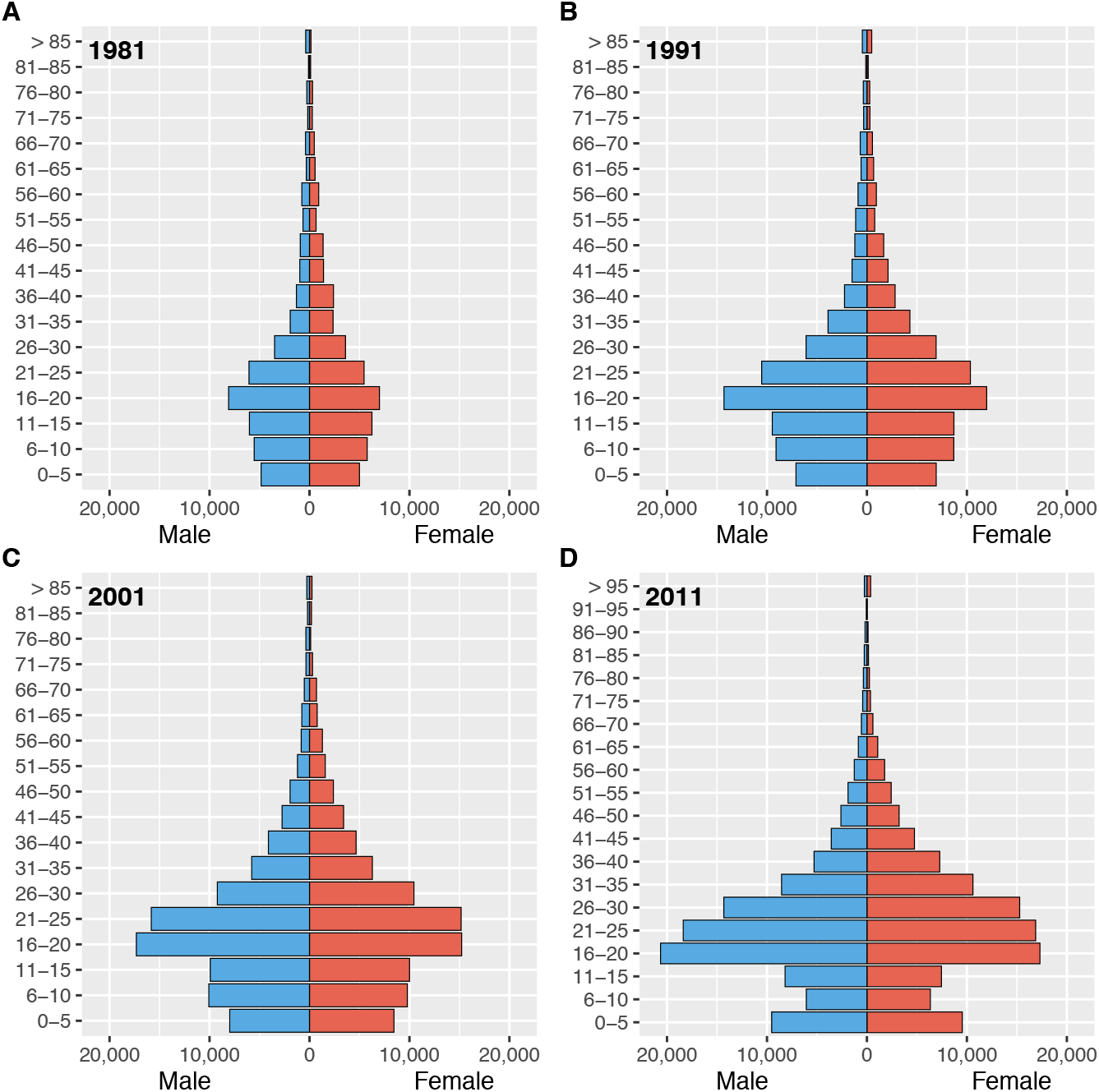
Gender-stratified age profiles of internal migrants by census year. **(A)** 1981, **(B)** 1991, **(C)** 2001, and **(D)** 2011.

### Migratory flows

To reconstruct the internal migration networks, we first calculated migratory flows between districts. We define a district-level migratory flow as the number of migrants who changed their residency from one district to another during the 12 months prior to the census. The migratory flows for each district (within district, in-flow, and out-flow) are listed in Table 1 (1980-1981), Table 2 (1990-1991), Table 3 (2000-2001), and Table 4 (2010-2011). Over three decades (between 1981 and 2011), the number of migrants more than doubled (as did the population of Botswana).

**Table 1:** Migration metrics by district, 1980-81. The table presents each district’s urban/rural classification, population size, total number of migrants (both within and between districts), within district migration intensity per 100 residents (WDMI), and population turnover. The urban/rural classes were: city (C), town (T), or predominantly rural (R). Population sizes are tabulated for residents for whom there was one-year migration data. Turnover was calculated as the net change in the annual migration rate per 100 residents.

| ID | District | Class | Population | Number of Migrants |  |  | Metric |  |
| --- | --- | --- | --- | --- | --- | --- | --- | --- |
|  |  |  |  | Within | In | Out | WDMI | Turnover |
| 1 | Gaborone | C | 56,390 | 120 | 7,750 | 6,110 | 0.22 | 3.00 |
| 2 | Francistown | C | 31,090 | 50 | 4,730 | 3,880 | 0.17 | 2.81 |
| 3 | Lobatse | T | 18,110 | 0 | 1,940 | 2,260 | 0.00 | -1.74 |
| 4 | Selebi Phikwe | T | 27,210 | 50 | 4,080 | 3,850 | 0.19 | 0.85 |
| 5 | Orapa | T | 6,330 | 0 | 810 | 890 | 0.00 | -1.25 |
| 6 | Jwaneng | T | 5,120 | 0 | 2,310 | 710 | 0.00 | 45.45 |
| 10 | Southern | R | 103,190 | 7,120 | 3,330 | 3,390 | 6.90 | -0.06 |
| 11 | Barolong | R | 14,290 | 170 | 340 | 770 | 1.15 | -2.92 |
| 20 | South East | R | 30,880 | 330 | 2,000 | 1,000 | 1.10 | 3.35 |
| 30 | Kweneng | R | 109,890 | 5,970 | 2,010 | 2,450 | 5.41 | -0.40 |
| 40 | Kgatleng | R | 43,110 | 1,740 | 1,330 | 1,960 | 3.98 | -1.44 |
| 50 | Central Serowe | R | 93,260 | 5,240 | 3,350 | 4,480 | 5.55 | -1.20 |
| 51 | Central Mahalapye | R | 78,910 | 3,120 | 2,530 | 3,200 | 3.92 | -0.84 |
| 52 | Central Bobonong | R | 44,160 | 3,100 | 1,840 | 2,730 | 6.88 | -1.98 |
| 53 | Central Boteti | R | 24,620 | 1,940 | 1,200 | 1,050 | 7.93 | 0.61 |
| 54 | Central Tutume | R | 74,850 | 2,790 | 3,230 | 3,940 | 3.69 | -0.94 |
| 55 | Central Tuli Block | R | 2,740 | 20 | 530 | 0 | 0.90 | 23.98 |
| 60 | North East | R | 36,360 | 890 | 1,890 | 2,480 | 2.41 | -1.60 |
| 70 | Ngamiland South | R | 58,160 | 2,870 | 1,280 | 1,360 | 4.93 | -0.14 |
| 71 | Ngamiland North | R | 7,760 | 90 | 140 | 310 | 1.13 | -2.14 |
| 72 | Chobe | R | 7,390 | 480 | 370 | 380 | 6.49 | -0.14 |
| 73 | Okavango Delta | R | 1,020 | 20 | 150 | 20 | 2.25 | 14.61 |
| 80 | Ghanzi | R | 17,890 | 1,620 | 390 | 530 | 8.99 | -0.78 |
| 90 | Kgalagadi South | R | 14,910 | 690 | 450 | 420 | 4.64 | 0.20 |
| 91 | Kgalagadi North | R | 8,880 | 170 | 430 | 240 | 1.96 | 2.19 |
| Botswana |  |  | 916,520 | 38,590 | 48,410 | 48,410 | 4.21 | 0 |

**Table 2:** Migration metrics by district, 1990-91. The table presents each district’s urban/rural classification, population size, total number of migrants (both within and between districts), within district migration intensity per 100 residents (WDMI), and population turnover. The urban/rural classes were: city (C), town (T), predominantly urban (U), partially urban (PU) and predominantly rural (R). Population sizes are tabulated for residents for whom there was one-year migration data. Turnover was calculated as the net change in the annual migration rate per 100 residents.

| ID | District | Class | Population | Number of Migrants |  |  | Metric |  |
| --- | --- | --- | --- | --- | --- | --- | --- | --- |
|  |  |  |  | Within | In | Out | WDMI | Turnover |
| 1 | Gaborone | C | 137,680 | 550 | 15,350 | 14,620 | 0.40 | 0.53 |
| 2 | Francistown | C | 67,740 | 70 | 8,110 | 6,510 | 0.11 | 2.42 |
| 3 | Lobatse | T | 26,570 | 0 | 3,130 | 2,640 | 0.00 | 1.88 |
| 4 | Selebi Phikwe | T | 40,480 | 30 | 4,920 | 4,110 | 0.08 | 2.04 |
| 5 | Orapa | T | 8,120 | 0 | 900 | 1,080 | 0.00 | -2.17 |
| 6 | Jwaneng | T | 10,520 | 0 | 1,450 | 1,800 | 0.00 | -3.22 |
| 7 | Sowa | T | 1,640 | 0 | 780 | 410 | 0.00 | 29.13 |
| 10 | Southern | R | 125,400 | 5,200 | 5,020 | 6,460 | 4.10 | -1.14 |
| 11 | Barolong | R | 18,050 | 420 | 700 | 1,400 | 2.24 | -3.73 |
| 20 | South East | U | 42,320 | 900 | 3,280 | 2,150 | 2.18 | 2.74 |
| 31 | Kweneng East | PU | 140,810 | 6,650 | 8,060 | 6,480 | 4.78 | 1.13 |
| 32 | Kweneng West | R | 27,700 | 1,360 | 1,630 | 1,430 | 4.95 | 0.73 |
| 40 | Kgatleng | PU | 55,180 | 2,670 | 3,040 | 3,200 | 4.82 | -0.29 |
| 50 | Central Serowe | R | 126,810 | 7,050 | 7,350 | 6,900 | 5.58 | 0.36 |
| 51 | Central Mahalapye | R | 97,030 | 5,360 | 4,690 | 6,360 | 5.43 | -1.69 |
| 52 | Central Bobonong | R | 51,220 | 2,780 | 2,220 | 3,320 | 5.31 | -2.10 |
| 53 | Central Boteti | R | 37,630 | 2,190 | 1,980 | 1,770 | 5.85 | 0.56 |
| 54 | Central Tutume | R | 98,940 | 3,590 | 5,000 | 6,960 | 3.56 | -1.94 |
| 60 | North East | R | 41,470 | 1,040 | 2,830 | 3,290 | 2.48 | -1.10 |
| 70 | Ngamiland South | PU | 60,450 | 4,650 | 2,470 | 4,400 | 7.45 | -3.09 |
| 71 | Ngamiland North | R | 33,750 | 620 | 2,650 | 650 | 1.95 | 6.30 |
| 72 | Chobe | PU | 12,500 | 700 | 1,320 | 1,100 | 5.70 | 1.79 |
| 80 | Ghanzi | R | 26,110 | 1,970 | 820 | 810 | 7.55 | 0.04 |
| 90 | Kgalagadi South | R | 19,250 | 1,010 | 910 | 780 | 5.28 | 0.68 |
| 91 | Kgalagadi North | R | 10,970 | 410 | 740 | 720 | 3.74 | 0.18 |
| Botswana (Total) |  |  | 1,318,340 | 49,220 | 89,350 | 89,350 | 3.73 | 0 |

**Table 3:** Migration metrics by district, 2000-01. The table presents each district’s urban/rural classification, population size, total number of migrants (both within and between districts), within district migration intensity per 100 residents (WDMI), and population turnover. The urban/rural classes were: city (C), town (T), predominantly urban (U), partially urban (PU) and predominantly rural (R). Population sizes are tabulated for residents for whom there was one-year migration data. Turnover was calculated as the net change in the annual migration rate per 100 residents.

| ID | District | Class | Population | Number of Migrants |  |  | Metric |  |
| --- | --- | --- | --- | --- | --- | --- | --- | --- |
|  |  |  |  | Within | In | Out | WDMI | Turnover |
| 1 | Gaborone | C | 195,150 | 1,360 | 22,100 | 25,420 | 0.69 | -1.67 |
| 2 | Francistown | C | 83,900 | 220 | 9,060 | 9,620 | 0.26 | -0.66 |
| 3 | Lobatse | T | 31,510 | 170 | 3,660 | 4,120 | 0.53 | -1.44 |
| 4 | Selebi Phikwe | T | 50,420 | 110 | 5,420 | 5,630 | 0.22 | -0.41 |
| 5 | Orapa | T | 8,770 | 10 | 1,450 | 1,100 | 0.12 | 4.16 |
| 6 | Jwaneng | T | 15,360 | 70 | 2,870 | 2,330 | 0.47 | 3.64 |
| 7 | Sowa | T | 3,150 | 0 | 680 | 790 | 0.00 | -3.37 |
| 10 | Southern | R | 113,600 | 4,850 | 6,940 | 7,670 | 4.24 | -0.64 |
| 11 | Barolong | R | 45,450 | 1,310 | 3,310 | 3,070 | 2.90 | 0.53 |
| 12 | Ngwaketse West | U | 10,840 | 380 | 930 | 1,000 | 3.48 | -0.64 |
| 20 | South East | U | 59,670 | 690 | 6,030 | 3,680 | 1.20 | 4.10 |
| 31 | Kweneng East | U | 185,880 | 6,010 | 11,760 | 9,540 | 3.27 | 1.21 |
| 32 | Kweneng West | R | 39,110 | 2,040 | 2,220 | 2,540 | 5.17 | -0.81 |
| 40 | Kgatleng | PU | 73,460 | 2,270 | 4,250 | 4,090 | 3.10 | 0.22 |
| 50 | Central Serowe | PU | 151,830 | 5,750 | 9,830 | 10,780 | 3.76 | -0.62 |
| 51 | Central Mahalapye | PU | 110,010 | 4,560 | 6,250 | 6,980 | 4.12 | -0.66 |
| 52 | Central Bobonong | PU | 64,330 | 2,710 | 4,090 | 3,420 | 4.26 | 1.05 |
| 53 | Central Boteti | PU | 47,110 | 3,780 | 3,110 | 2,140 | 8.19 | 2.10 |
| 54 | Central Tutume | R | 121,520 | 4,390 | 6,630 | 8,030 | 3.57 | -1.14 |
| 60 | North East | R | 49,930 | 1,310 | 3,740 | 3,630 | 2.63 | 0.22 |
| 70 | Ngamiland South | U | 69,370 | 2,350 | 4,890 | 4,320 | 3.42 | 0.83 |
| 71 | Ngamiland North | R | 52,820 | 2,550 | 2,130 | 2,190 | 4.82 | -0.11 |
| 72 | Chobe | PU | 16,250 | 750 | 1,700 | 990 | 4.83 | 4.57 |
| 80 | Ghanzi | R | 32,370 | 3,090 | 1,570 | 2,030 | 9.41 | -1.40 |
| 90 | Kgalagadi South | R | 25,260 | 1,680 | 1,400 | 1,420 | 6.65 | -0.08 |
| 91 | Kgalagadi North | R | 16,510 | 910 | 1,700 | 1,190 | 5.69 | 3.19 |
| Botswana (Total) |  |  | 1,673,580 | 53,320 | 127,720 | 127,720 | 3.19 | 0 |

**Table 4:**
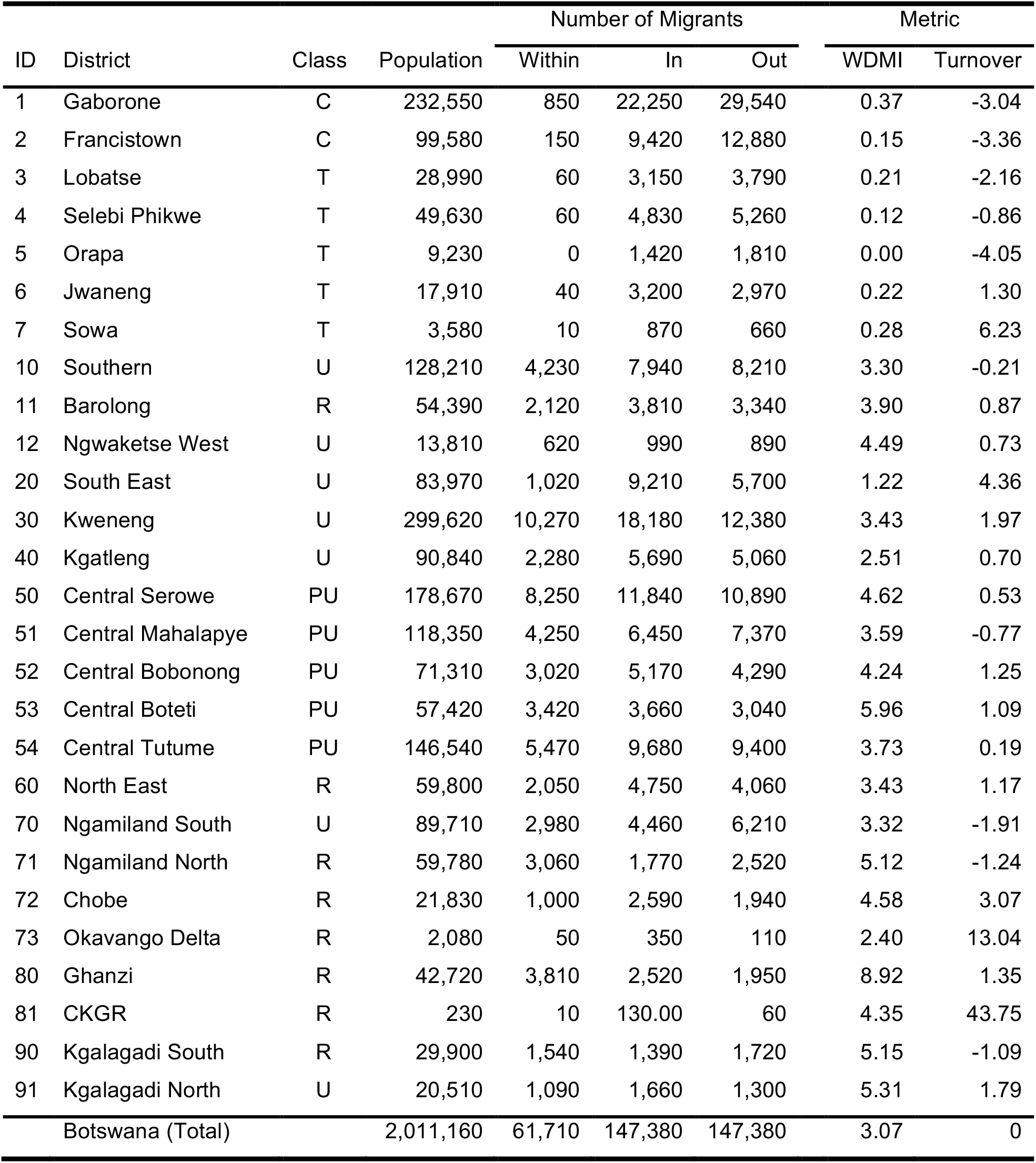
Migration metrics by district, 2010-11. The table presents each district’s urban/rural classification, population size, total number of migrants (both within and between districts), within district migration intensity per 100 residents (WDMI), and population turnover. The urban/rural classes were: city (C), town (T), predominantly urban (U), partially urban (PU) and predominantly rural (R). Population sizes are tabulated for residents for whom there was one-year migration data. Turnover was calculated as the net change in the annual migration rate per 100 residents. The Central Kgalagadi Game Reserve district is denoted CKGR.

In 1981, the two cities and four towns were fairly small. The capital, Gaborone, had a population of 56,860 and Francistown had a population of only 31,120. The populations of the four towns ranged in size from 5,200 to 27,430. The number of individuals who lived in the other districts ranged in size from 1,050 in the Okavango Delta to 111,820 in Kweneng. All of these districts had an in-flow, and/or out-flow, of migrants (Table 1). More than half also had a fairly high within district migration intensity (WDMI): for example, in Central Bobonong, ∼7% of residents moved from one rural village to another between 1980 and 1981. Migratory flows continued to increase over the next three decades (Tables 2-4), as the population size increased and the country became progressively urbanized.

We also estimated the annual turnover rate in each district in the year before each census (Tables 1-4). This rate is defined as the net change in the district’s annual migration rate per hundred residents. A negative rate signifies that the district’s population size decreased, a positive rate signifies that it increased. By far, the highest turnover rates were in 1981 (Table 1). In that year, the highest turnover rate was in the mining town, Jwaneng.

### Reconstructed internal migration networks

The chord diagrams (Figure 2) show the reconstructed internal migration networks, based on the micro-census data, between: 1980-81 (Figure 2A), 1990-91 (Figure 2B), 2000-01 (Figure 2C), and 2010-11 (Figure 2D). Each district is a node in the network. Networks are shown in terms of the magnitude of the migratory in-flows and out-flows between districts, and the effect of these flows on connecting different districts throughout the country: two districts are connected if they have a migratory flow between them. Even the earliest migration network in 1981 – when Botswana was predominantly rural – can be seen to be fairly complex and shows a high degree of connectivity amongst the districts, although the migratory flows and counter-flows are fairly small. Notably, in all four networks, many of the flows and counter-flows are similar in magnitude. Both the magnitude of the migratory flows, and counter-flows, increased with time (between 1981 and 2011) as the population increased in size.

**Figure 2:**
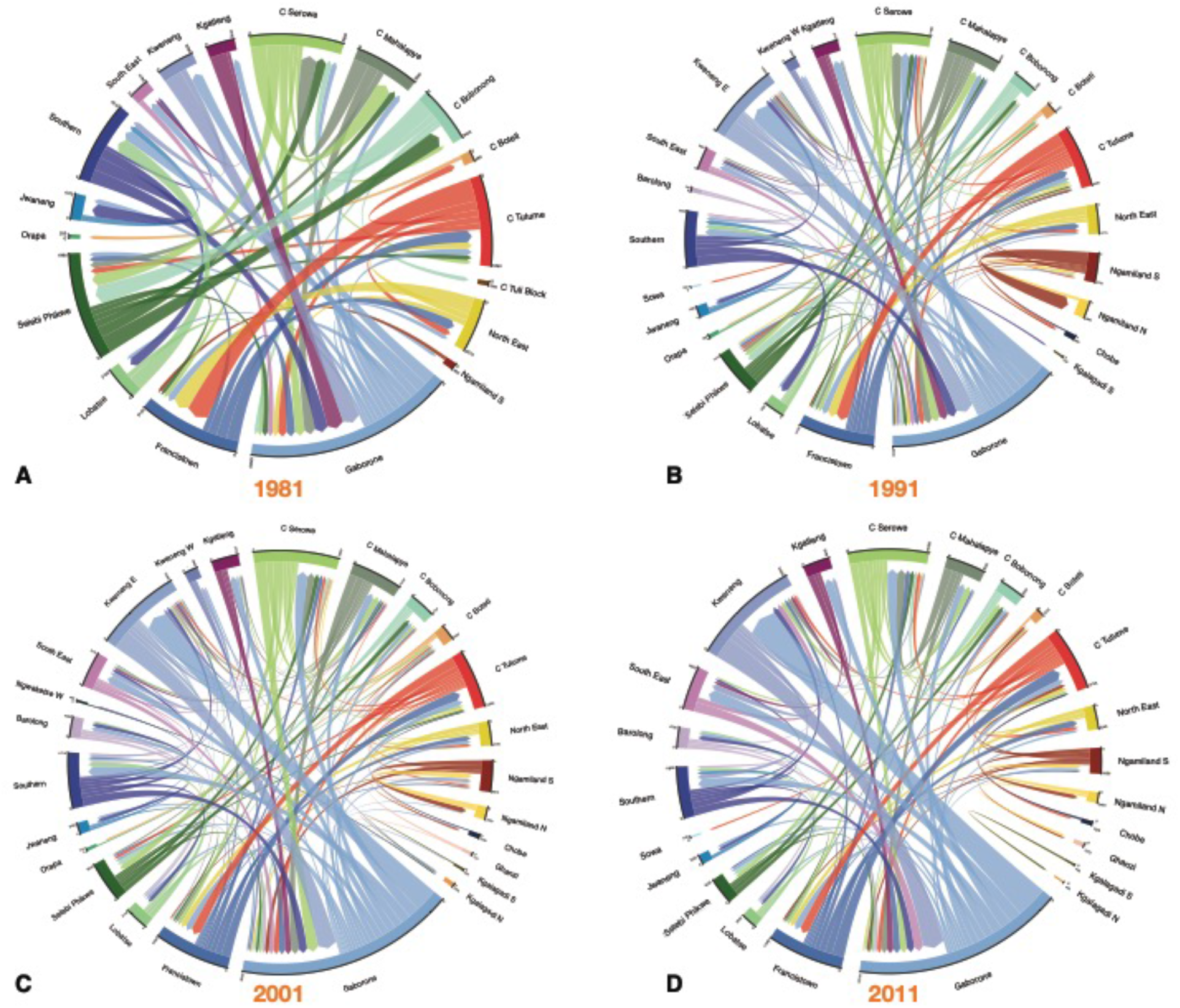
Chord diagrams by census year. **(A)** 1981, **(B)** 1991, **(C)** 2001, and **(D)** 2011. Each diagram shows the internal migration network of the general population in the 12 months prior to the census. Each color represents a different administrative district. The thickness of each line is proportional to the number of migrants that moved between the two connected districts. The angular width of each district is proportional to the total number of migrants who moved into, or out of, that district. For clarity, in (A)-(C) only connections with greater than 200 migrants are shown, and in (D) only connections with greater than 400 migrants are shown. Consequently, some districts are not shown in the chord diagram. The total number of migrants (in and out) of every district is listed in Tables 1-4.

### In-flow and out-flow migration hubs

Out-flow and in-flow migration hubs are shown in Figure 3. Notably, the towns (Jwaneng, Selebi Phikwe, Sowa, Lobatse, and Orapa) were amongst the top five out-flow and in-flow migration hubs at every census period.

**Figure 3:**
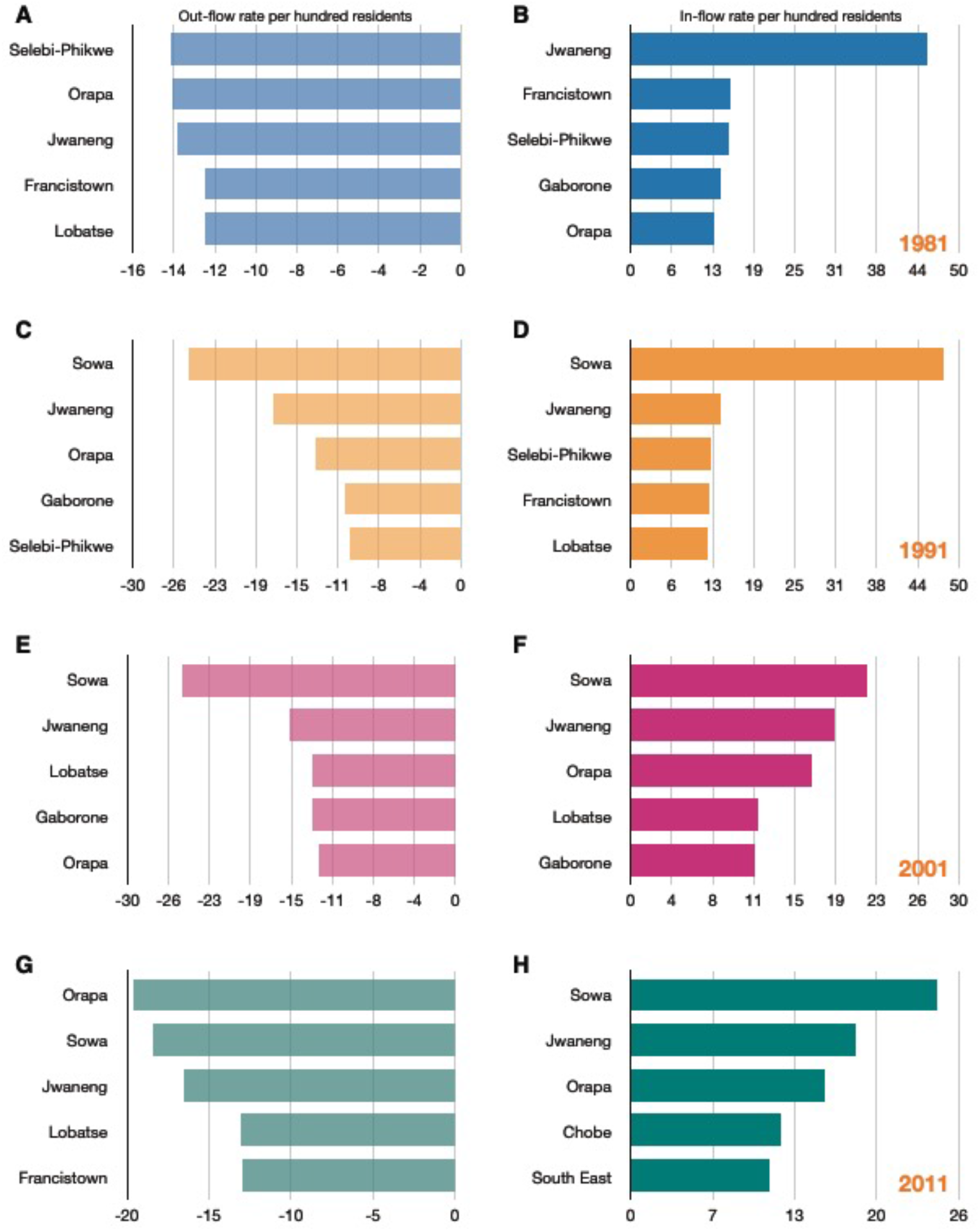
The top five out-flow and in-flow migration hubs by census year. **(A)** Top out-flow hubs in 1981, **(B)** Top in-flow hubs in 1981, **(C)** Top out-flow hubs in 1991, **(D)** Top in-flow hubs in 1991, **(E)** Top out-flow hubs in 2001, **(F)** Top in-flow hubs in 2001, **(G)** Top out-flow hubs in 2011, **(H)** Top in-flow hubs in 2011. Out-flow hubs are sorted by the rate per hundred residents of a district’s population that moves out to another district; in-flow hubs are sorted by the rate per hundred residents of a district’s population that moves in from another district.

In 1981, the top in-flow hub was the diamond mining town, Jwaneng: 45% of the town’s population consisted of migrants who had moved to the town between 1980 and 1981 (Figure 3B). At that time, Jwaneng was also an important out-flow hub: 14% of the town’s population moved to another district between 1980 and 1981 (Figure 3A). The ego networks for Jwaneng show the districts that migrants returned to (Figure 4A), and the districts that they came from (Figure 4B). It can be seen that migrants moved between the other mining towns, the two cities, and many of the rural districts, with the majority of migrants moving between Jwaneng and Southern, a district abutting the town (Figure 2—figure supplement 1).

**Figure 4:**
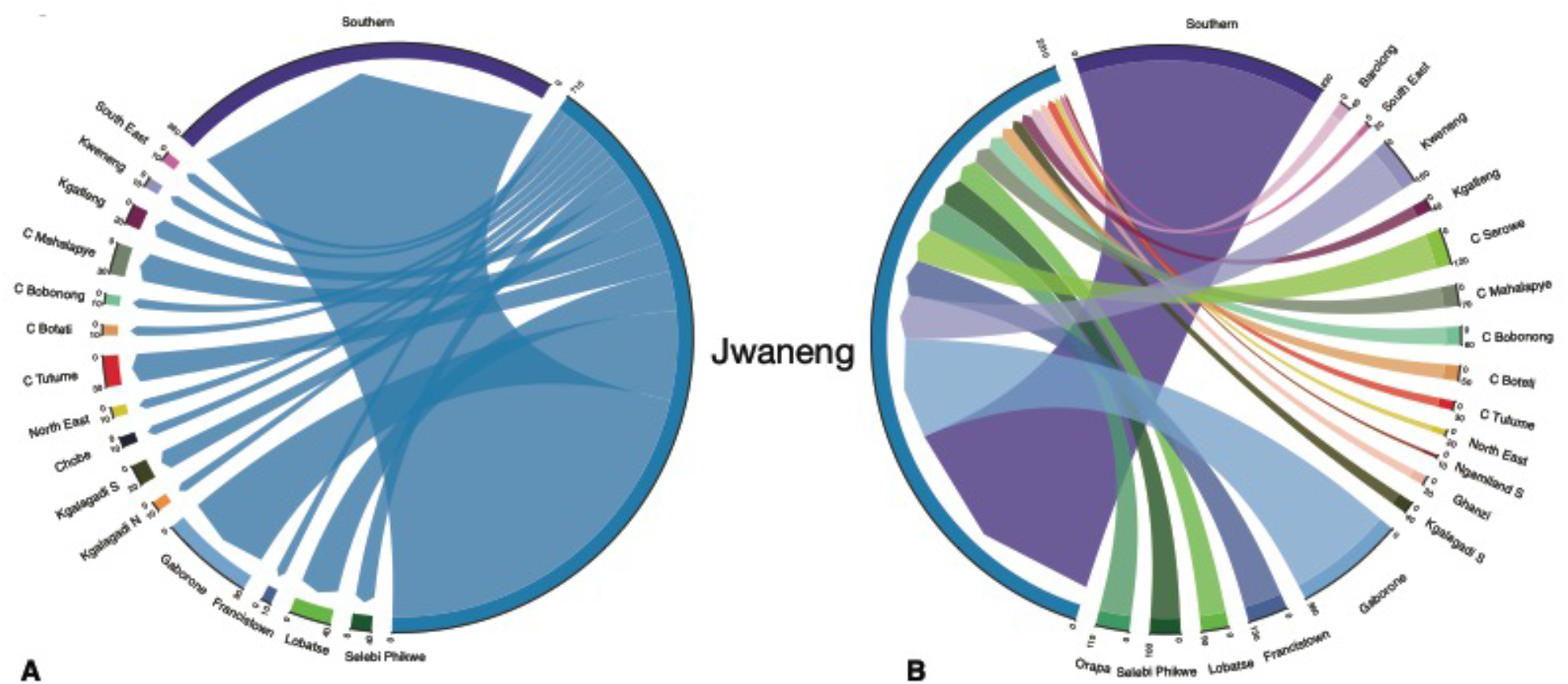
Ego network of Jwaneng. The chord diagrams show migrants flowing **(A)** out of, and **(B)** into, the diamond mining town in 1981.

In 1991 and 2001, the top out-flow (and in-flow) hub was the mining town, Sowa. In 1991, 48% of the population consisted of migrants who had moved to the town in the previous 12 months (Figure 3D); 25% of the town’s population moved to another district between 1990 and 1991 (Figure 3C). In 2001, 22% of the population consisted of migrants who had moved to the town in the previous 12 months (Figure 3F); 25% of the town’s population moved to another district in 2000-2001 (Figure 3E).

Francistown was an important in-flow and out-flow hub between 1980 and 1981. In 1981, 15% of the population consisted of migrants who had moved to the town in the previous 12 months (Figure 3B); 13% of the town’s population moved to another district between 1980 and 1981 (Figure 3A). The city was also an important in-flow hub in 1991 (Figure 3D) and out-flow hub in 2011 (Figure 3G). Gaborone was only an important out-flow hub in 1991 and 2001 (Figure 3C and Figure 3E); it was also an important in-flow hub in 1981 and 2001 (Figure 3B and Figure 3F).

### Quantifying urban-to-rural and rural-to-urban migratory flows

The internal migration networks for each of the four time periods (1981, 1991, 2001, and 2011) are presented in terms of our urban/rural classification framework, both as schemata (Figure 5) and as Sankey diagrams (Figure 6). Both the schemata and the Sankey diagrams show the magnitude of the migratory flows amongst the five classes in the 12 months before each census. Between 1981 and 2011 the overall percentage of the population living in urban areas increased more than 3.5 fold (from ∼18% to ∼64%) (Statistics Botswana, 2014), and the internal migration networks changed substantially over that period.

**Figure 5:**
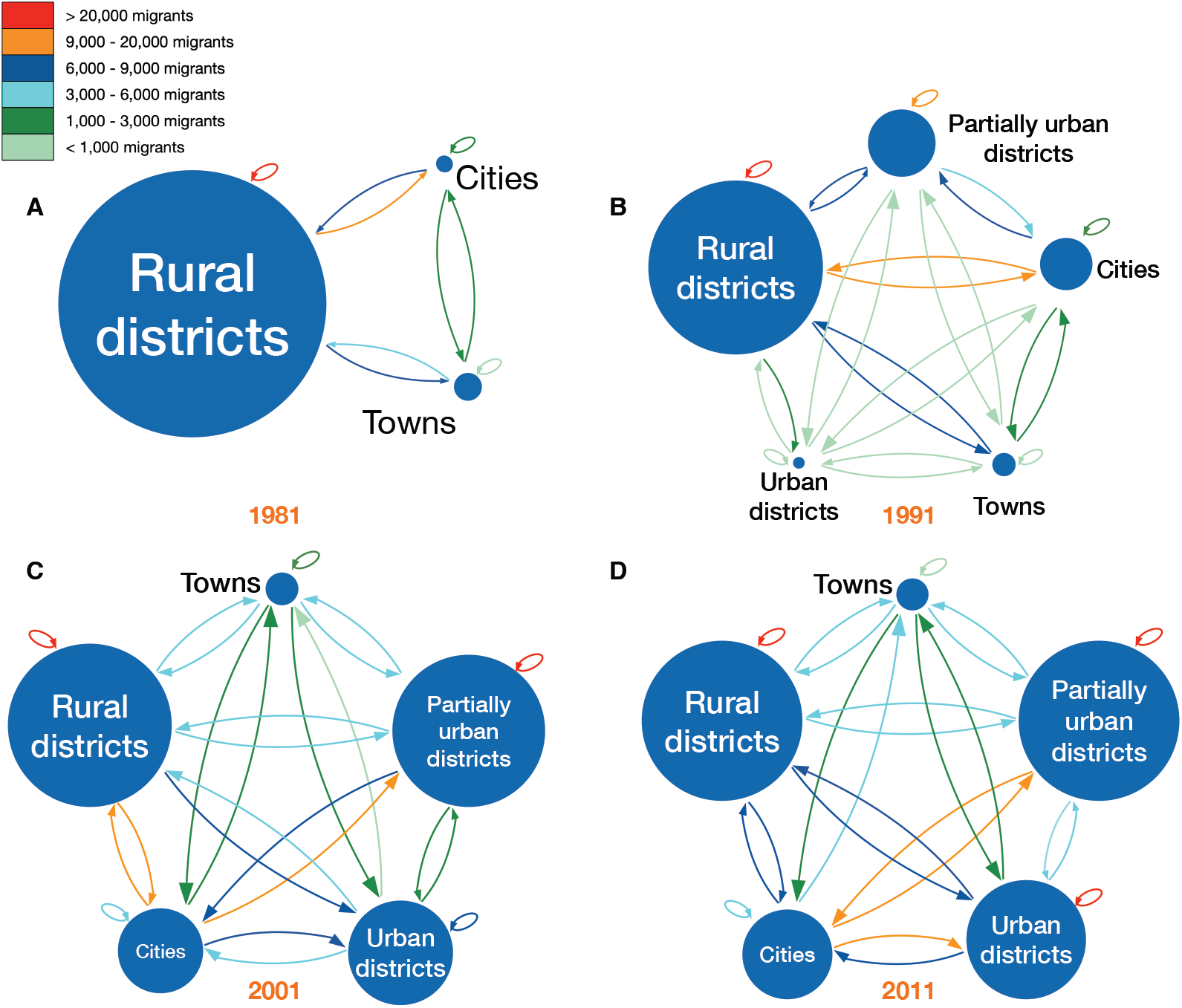
Schemata of urban/rural classification framework by census year. **(A)** 1981, **(B)** 1991, **(C)** 2001, and **(D)** 2011. Circles represent the five classes based on an urban/rural classification (see Methods). The radius of each circle is proportional to the number of residents living in the districts in that specific class. The color of each arrow indicates the size of the net migration between classes. The class designation (city, town, predominantly urban, partially urban, predominantly rural) of each district over time is provided in Tables 1-4.

**Figure 6:**
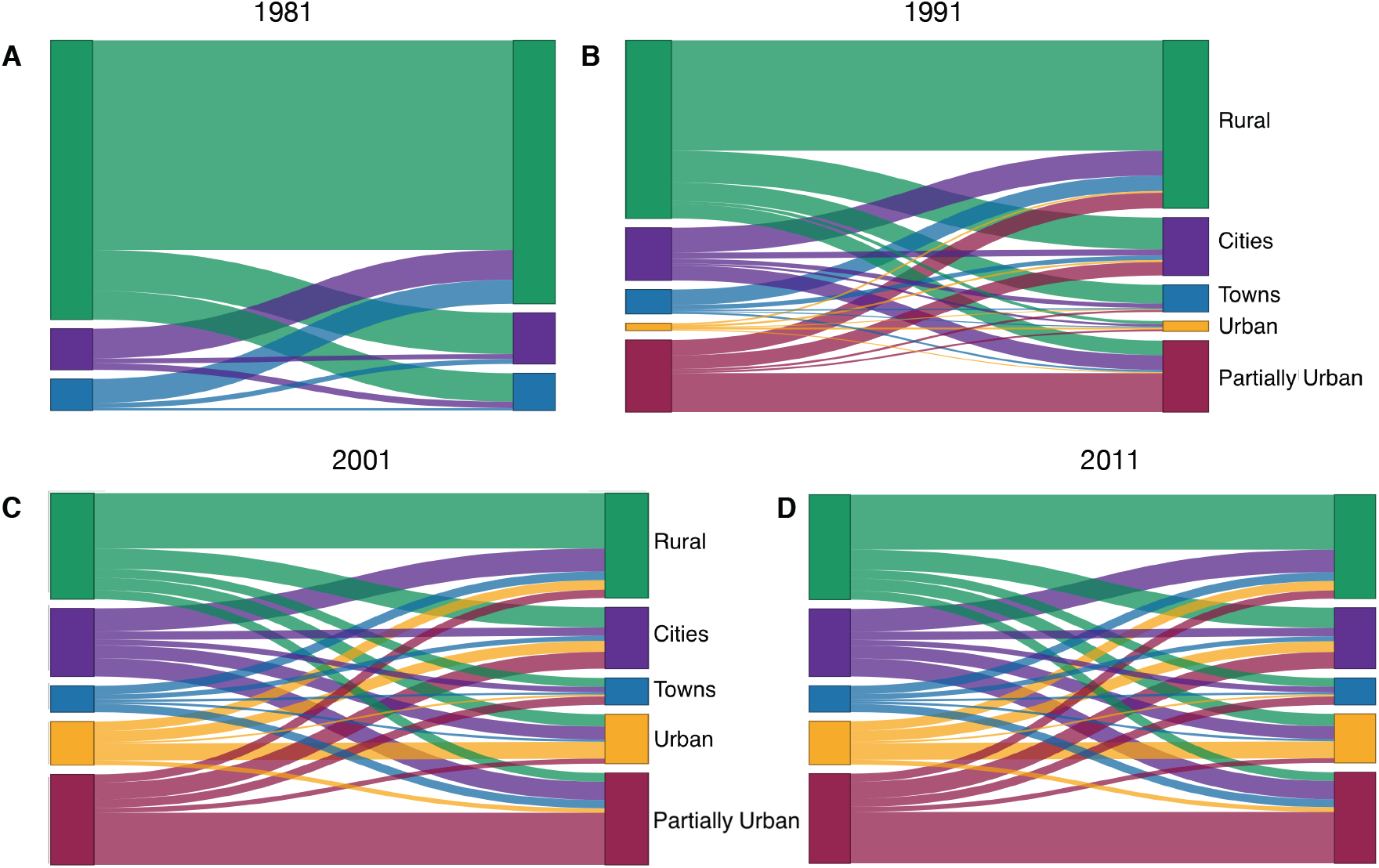
Sankey diagrams showing urban/rural classification framework by census year. **(A)** 1981, **(B)** 1991, **(C)** 2001, and **(D)** 2011. The diagrams show the relative magnitude of migratory flows between the different classes over time. Cities are shown in purple, towns in blue, predominantly urban districts in gold, partially urban districts in magenta, and predominantly rural districts in green. The class designation (city, town, predominantly urban, partially urban, predominantly rural) of each district over time is provided in Tables 1-4.

In 1981, there were only six districts (the two cities and four towns) that were urban areas (Table 1, Figure 5A, and Figure 6A). The other districts contained several hundred small rural villages and no urbanized areas. Almost all districts had an in-flow and out-flow of migrants: notably, rural-to-urban, and counter flows (urban-to-rural) are apparent. Taken together the results show that in 1981 the population was very mobile, the majority of migratory flows were within and amongst rural districts, and rural-to-urban migrations were greater than urban-to-rural counter-flows.

By 1991, due to in situ urbanization, one previously rural district (South East) had become predominantly urban, and four districts (Kweneng East, Kgatleng, Ngamiland South, and Chobe) had become partially urban (Table 2). Migratory flows appear to be fairly symmetrical (in terms of the number of migrants who moved between pairs of districts) throughout the country (Figure 5B and Figure 6B). The towns had increased ∼1.5 fold since 1981, and the cities had more than doubled in size. As at the time of the previous census, there were considerable rural-to-urban and urban-to-rural migratory flows.

By 2001, considerably more in situ urbanization had occurred: since 1981, four districts had become predominantly urban, six districts partially urban, and only nine districts remained predominantly rural (Table 3). Notably, some of the migratory flows were now asymmetric: the number of migrants moving from the cities to the predominantly urban and partially urban districts were greater than the counter-flows (Figure 5C and Figure 6C). The growth rate of towns remained approximately the same as in the previous decade, but the growth rate of the cities had decreased to ∼1.4 fold; these rates depended upon the number of migrants, births, and deaths.

By 2011, twelve districts had become predominantly or partially urbanized (Table 4). The growth rate of cities had decreased to ∼1.2 fold, and the towns had not increased substantially in size. At this time, while most flows were symmetric, migrants were disproportionately leaving the cities in favor of towns and urban villages (Figure 5D and Figure 6D). Approximately 42% of the population were living in these urban villages; there was at least one urban village in every district.

## DISCUSSION

Our results support our hypothesis that – during the development of Botswana’s generalized HIV epidemic (i.e., between 1981 and 2011) – there was a high level of population mobility, and substantial urban-to-rural migratory flows. Using our time series of historical data, we found ∼10% of the population moved their residency in the 12 months before each census in 1981, 1991, 2001, and 2011. The constancy of this value at each census suggests that the annual migration rate in Botswana was stable at ∼10% for each year between 1981 and 2011. Notably, the type of migrants also remained constant over this time period: evenly split by gender, with younger people more likely to migrate. We found that migration occurred within districts, as well as between districts, but that migration between districts was more common. Even in 1981, at the beginning of the period of rapid urbanization, the migration network was highly connected and therefore large-scale population movements linked almost all of the districts in the country. Notably, we found at each census, that although the number of urbanized districts increased (as did their population size) the vast majority of migratory flows between the rural and developing urban districts were balanced by counter-flows.

Our results provide insights into how HIV could have diffused through the population of Botswana as the epidemic became generalized. The first cases of AIDS in Botswana were reported in the 1980s (African Natural Resources Center, 2016); this suggests that HIV may have been circulating in Botswana in the 1970s, as HIV has an incubation period of 10 to 12 years (Hendriks et al., 1993; Muñoz et al., 1989). One phylogenetic study indicates that HIV was introduced into Botswana in the 1960s, but exponential growth did not begin until the 1980s (Wilkinson et al., 2015). Another study suggests that HIV spread southwards from Kinshasa (the epicenter of the HIV epidemic) in the Democratic Republic of the Congo via travelers on the railroad that was built in the 1800s (Faria et al., 2014); notably, this railroad goes through Francistown. It is possible that HIV was introduced into the mining towns of Botswana from overseas or South Africa. In the 1970s and 1980s many young men from Botswana went to work in the mines in South Africa, returning as the mining industry became established in their home country. Our results show that, by 1981, the migration networks (for women and men) linked almost every district in Botswana; this indicates that residents from Francistown (UNAIDS & World Health Organization, 2004) and the mining towns (Barnett et al., 2002) could have seeded sub-epidemics in multiple rural districts throughout the country. HIV could then have spread widely within these districts due to the high rates of within-district migration. Due to the high migration rates, the sub-epidemics in rural districts were likely to have been seeded numerous times. When HIV was first introduced into rural districts, the sub-epidemics in these districts may have been maintained by source-sink dynamics (Okano et al., 2020): districts where transmission was high enough to be self-sustaining (e.g., mining towns) could have maintained sub-epidemics in rural districts where transmission was too low to be self-sustaining. Our results suggest that migratory flows between high and low prevalence areas are likely to have existed, at least from the early 1980s onwards. These flows would have functioned as “transmission corridors” (Okano et al., 2021), and created high risk flows (Valdano et al., 2021). Taken together, our quantitative results show that the highly-connected migration networks, with high migratory flows and increasing urbanization, could have contributed to the development of Botswana’s generalized epidemic.

Our results also provide insights into how the Botswana epidemic could have become hyper-endemic. Migrants have been shown to be at high risk of acquiring HV infection (Camlin & Charlebois, 2019; Dobra et al., 2017; Olawore et al., 2018); our results suggest that, due to migration alone, each year ∼10% of the population of Botswana was at high risk. The migrants in Botswana were young women and men, a group generally at the highest risk of infection through sexual transmission. Mining towns and cities would have been transmission hot-spots (for both women and men) due to high levels of risky sexual behavior (as evidenced by high prevalence (Barnett et al., 2002)) and a high probability of encountering an HIV-infected individual as a sex partner. Due to the high prevalence (Barnett et al., 2002; UNAIDS & World Health Organization, 2004), high turnover rates, and a high level of connectivity to other districts, the mining towns and cities could have functioned as geographically-defined core groups for sexual transmission (for both women and men). Some individuals could have been part of a high-risk core group when they were living in the cities and mining towns, and decreased their risky behavior when they returned to the rural districts. For example, FSWs in Botswana can have an extremely high number of sex partners in mining towns and urban areas (∼7 per week) and remain as FSWs for ∼4 years (Ministry of Health Botswana, 2013). Notably we found that the mining towns and cities were both migration in-flow hubs and migration out-flow hubs. The fact that they were migration in-flow hubs would have led to a high in-flow of uninfected individuals from rural districts; the fact that they were migration out-flow hubs would have led to a high out-flow of HIV-infected individuals to rural districts. Therefore, rural-to-urban migration coupled with urban-to-rural migration could have been a very important driver in the Botswana epidemic becoming hyper-endemic. Without this driver, prevalence could have reached very high levels in the mining towns and cities, but remained fairly low in rural districts.

Our study has several limitations. First, we have examined only internal migration (i.e., migration that takes place within a country), and have not included international migration. However, for the three-decade time period that we have investigated, the number of international migrants to Botswana was much smaller than the number of internal migrants (Statistics Botswana, 2014; Statistics Botswana, 2015). Second, all of our analyses are focused on Botswana in order to analyze the impact of migration networks on a hyper-endemic epidemic; we have not evaluated migration networks in other African countries. We recommend that studies, such as we have conducted here, are conducted in these other countries by analyzing a time series of historical micro-census data, especially in the only other two countries in sub-Saharan Africa that have generalized hyper-endemic HIV epidemics: Eswatini and Lesotho. Third, a potential limitation is that the IPUMS data are samples rather than the complete censuses; we used this approach as it is generally not possible to obtain complete census data due to privacy reasons. Fourth, while we have gained insights into understanding the development of the generalized hyper-endemic HIV epidemic in Botswana, we have not conducted an analysis of the transmission dynamics of the epidemic in order to explore these insights. This is the focus of current research in which we are analyzing a geospatial mathematical model of the epidemiological evolution of the HIV epidemic in Botswana. Fifth, we have examined only two processes that affected the evolution of Botswana’s epidemic, other factors could also have been important.

All HIV epidemics in SSA are generalized; HIV can only diffuse through a population by the movement of people. To our knowledge, our study is the first to study population-level mobility patterns and rates in any country in sub-Saharan Africa. Taken together, our results identify particular characteristics of migration in Botswana that could explain why HIV prevalence rose to very high levels in many districts throughout the country. Migratory behavior in other countries with generalized epidemics may be very different. For example, migrants in other African countries tend to have a very male-biased sex ratio; consequently, women are unlikely to have seeded multiple sub-epidemics throughout these countries. If mining towns and cities in these countries were not migratory in-flow and out-flow hubs with high turnover of their populations, as in Botswana, they would not have served as major sources of infection. Botswana has recently achieved the 95-95-95 targets that UNAIDS specified needed to be reached by 2030 in order to eliminate HIV: 95% of people living with HIV to be diagnosed, 95% of the diagnosed to be on treatment, and 95% of those on treatment to be virally suppressed (Mine et al., 2022). However, transmission is continuing (Magosi et al., 2022), as is urbanization and large-scale population movements (Okano et al., 2021). The same processes that we have identified that may have contributed to the development of the generalized hyper-endemic HIV epidemic in Botswana may prevent the elimination of HIV.

## MATERIALS AND METHODS

### Data

To conduct our analyses, we used representative samples of micro-census data extracted from the IPUMS-International (Integrated Public Use Microdata Series-International) database (Minnesota Population Center, 2020): micro-census data are anonymized individual-level data (Ruggles et al., 2015). IPUMS-International currently disseminates data from 547 censuses and surveys in 103 countries worldwide (Minnesota Population Center, 2020). The Botswana dataset consists of representative 10% samples from the original censuses, and includes anonymized individual-level data on age, gender, and residence (current, as well as 12 months prior). The data also includes individual survey weights that allow for population-level estimation (Ruggles et al., 2015).

To examine historical trends in internal migration networks and urbanization, we analyzed data collected in the 1981, 1991, 2001, and 2011 censuses. We used data on internal migration that had occurred in the 12 months prior to each census: an individual was classified as a migrant if they had changed their permanent residency within that 1-year interval.

### Estimating the incidence of internal migration

We estimated the incidence of internal migration by calculating the CMI. This statistic represents the overall incidence, or level of internal migration (between district plus within district), per hundred residents over a year. The mathematical definition of the CMI is given in Equation 1:

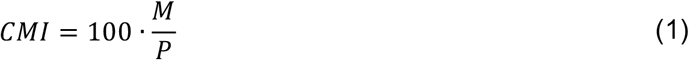

where *M* is the total number of internal migrants and *P* is the population size of Botswana in a given year. *M* is calculated as *M =* ∑_*i*_ *D*_*i*_ *=* ∑_*i*_ *O*_*i*_ where *D*_*i*_ represents the in-flows to each district *i*, and *O*_*i*_ represents the out-flows to each district *i*.

### Constructing gender-stratified age structure pyramids

We aggregated the migration data from each census by gender and age (using 5-year age groupings) to construct population pyramids; these pyramids show the age-gender demographics of all individuals internally migrating in the 12 months prior to each census.

### Calculating migratory flows

We define a district-level migratory flow as the number of migrants who change their residency from one district to another during the 12 months prior to the census. We calculated migratory flows between each pair of districts. The country consists of 28 administrative districts (Okano et al., 2021). Each city and town are separate administrative districts.

### Calculating annual turn-over rates

We defined the annual turn-over rate for a district as the net change in its annual migration rate per hundred residents.

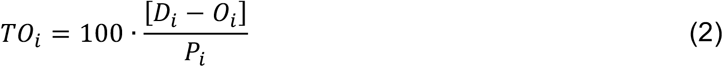

Here the turn-over rate *TO*_*i*_ for district *i* is a function of the number of in-migrants *D*_*i*_ and out-migrants *O*_*i*_ within the past year, and its population *P*_*i*_ at the beginning of the year.

### Reconstructing migration networks

We used the micro-census data to construct Origin-Destination (OD) matrices: the origin was the district that an individual lived in 12 months prior to the census, the destination was the district they lived in at the time of the census. Coefficients of these matrices specify the number of migrants who moved between each pair of districts in the 12 months prior to each census.

### Identifying in-flow and out-flow migration hubs

Migration hubs are those districts where recent migration has a sizeable impact on the size of the resident population, by either bringing it down (out-flow hubs) or increasing it (in-flow hubs) above the average. In-flow migration hubs were identified by calculating the total number of in-migrants to a node/district and dividing by the district’s population that year. Out-flow migration hubs were identified by calculating the total number of out-migrants to a node/district and dividing by the district’s population that year. We list the top five in-flow and out-flow hubs each census year.

### Quantifying urban-to-rural and rural-to-urban migratory flows

To measure and visualize migratory flows during urbanization, we developed a classification framework. This framework enables us to measure the magnitude of migratory flows and counter-flows between districts (based upon their degree of urbanization), and distinguishes between urban-to-rural and rural-to-urban flows.

The framework consists of five classes. The classes (at any point in time) are defined based on the degree of urbanization of the districts at the time of the most recent census: (i) predominantly rural (< 40% of the population living in urban areas), (ii) partially urban (40%-60% of the population living in urban areas), (iii) predominantly urban (> 60% of the population living in urban areas), (iv) town (100% of the population living in urban areas), or (v) city (100% of the population living in urban areas). Classes (i), (ii), and (iii) only include urban areas that develop by in situ urbanization, i.e., rural villages transforming into urban villages. There are migratory flows and counter-flows between the five classes, and flows within each of the five classes: therefore, 25 migratory flows/counter-flows are possible.

The classification framework can be visualized in two formats: (i) a schematic figure showing the five classes, the number of residents in each class, and migratory flows (and counter-flows) amongst and within the classes, and (ii) Sankey diagrams showing the number of migrants who move within, and between, the five classes.

We used the micro-census data to parameterize the framework for Botswana. Specifically, we estimated – at the time of each census – the population size of each district, and determined how many individuals lived in rural and urban areas. We used these estimates to classify all districts into one of the five classes. We then calculated the migratory flows and counter-flows between, and within, districts in the 12 months prior to each survey. These parameter estimates are given for 1981 census (Table 1), the 1991 census (Table 2), the 2001 census (Table 3), and the 2011 census (Table 4). Between each census, districts change in size due to births, deaths and migration. Also, between each census, districts can be reclassified based upon their increase in urbanization.

## Data Availability

All data needed to evaluate the conclusions in the paper are presented in the paper, or freely available for registered users at the IPUMS International website: https://doi.org/10.18128/D020.V7.3. We note that we did not collect these data, nor are they permitted to be posted to other repositories. All code needed to reproduce all parts of this analysis are available from the first author's GitHub page: https://github.com/janetsong80/pop-mobility-botswana-hiv (copy archived at: https://zenodo.org/badge/latestdoi/594401539).

https://doi.org/10.18128/D020.V7.3

## Acknowledgements

We acknowledge the Central Statistics Office (CSO) Botswana for collecting the data archived by IPUMS. We are grateful to Nelson Freimer for discussions throughout the course of this research.

## Author contributions

Conceptualization: LB, KS, SB

Methodology: JS, JTO, JP, EV

Software: JS, JP

Validation: JS, JTO

Formal analysis: JS, JTO, JP

Visualization: JTO, JP

Supervision: SB Writing—original draft: SB

Writing—review & editing: JS, JTO, JP, LB, KS, EV, SB

Funding acquisition: SB

---

**Figure 2—supplement 1:**
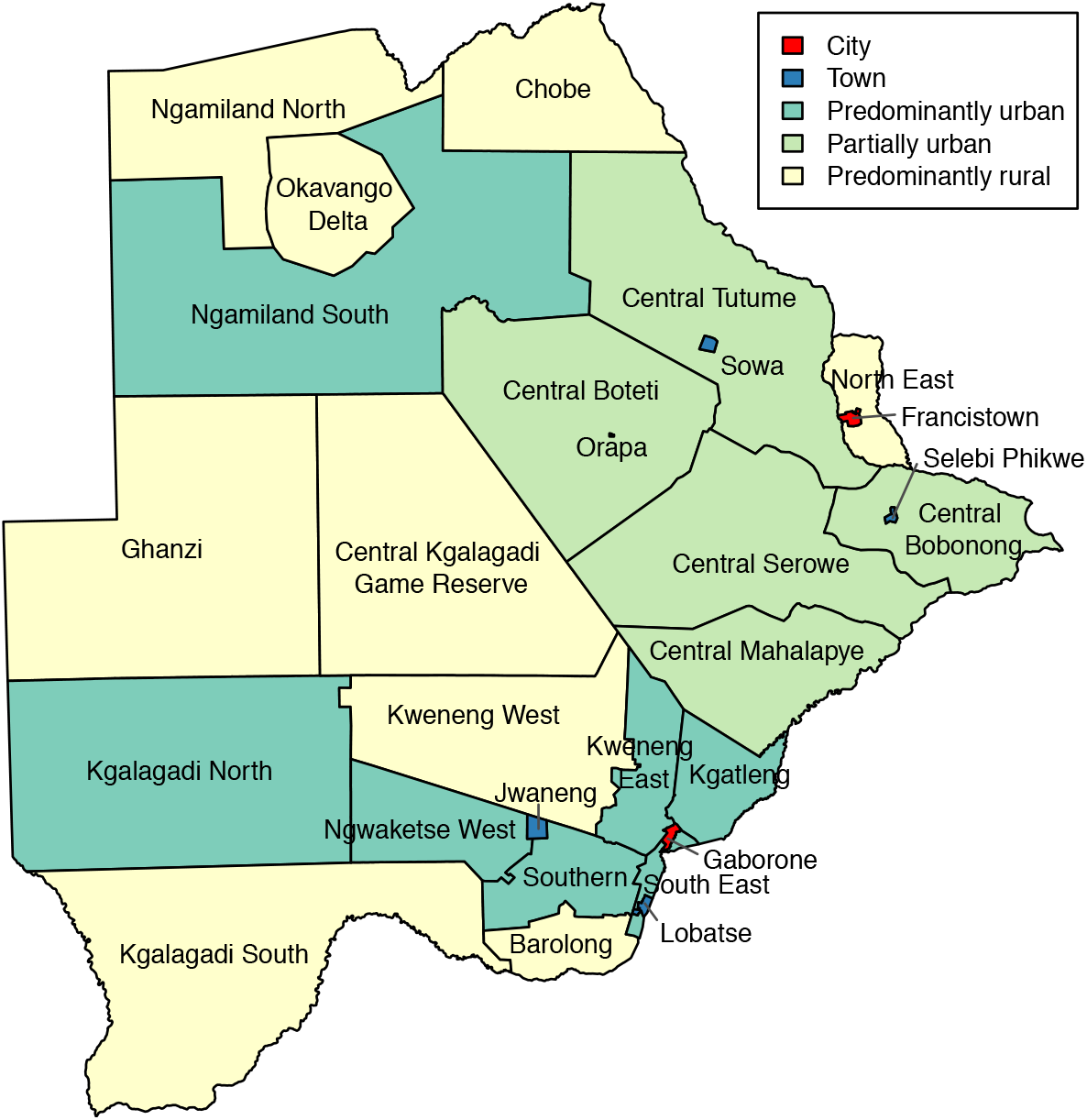
Map of Botswana with current district names, colored by class.

